# Prevalence of *dhfr-dhps* sextuple mutants and gametocyte-harboring quintuple mutants resistant to sulfadoxine-pyrimethamine among pregnant women in Mozambique

**DOI:** 10.64898/2026.03.31.26349751

**Authors:** Yasmina Drissi-El Boukili, Eduard Rovira-Vallbona, Pieter Guetens, Driss Chiheb, Johanna Helena Kattenberg, Luc Kestens, Sónia M. Enosse, Anna Rosanas-Urgell, Paulo Arnaldo

## Abstract

The intermittent preventive treatment with sulfadoxine-pyrimethamine (IPTp-SP) remains the main strategy to prevent malaria in pregnancy. However, continued drug pressure may also contribute to the emergence of resistant parasites and impact the gametocyte carriage and subsequent infectiousness. Pregnant women are thought to be a potential reservoir for malaria transmission due to the increased carriage of gametocytes following long-lasting infections. We used molecular methods to examine 100 *Plasmodium falciparum (P. falciparum*) isolates collected from Mozambican women at delivery in 2014-15, to determine SP resistance polymorphisms in *P. falciparum* dihydrofolate reductase (*pfdhfr*) and dihydropteroate synthetase (*pfdhps*) genes as well as the presence of gametocytes by RT-qPCR. Overall, 54% and 7% of parasites harbored quintuple and sextuple *pfdhfr/pfdhps* mutant haplotypes, respectively. Gametocytes were detected in 34% of isolates. Gametocyte carriage was significantly associated with quintuple mutant infections (AOR = 7.5, *p* = 0.001), which accounted for 80% of infections with detectable gametocytes. Results indicate the relevance of ongoing surveillance of SP resistance in Mozambique to guide future evaluation of alternative IPTp approaches as resistance levels evolve and to anticipate potential implications for parasite transmission and maternal-fetal health.

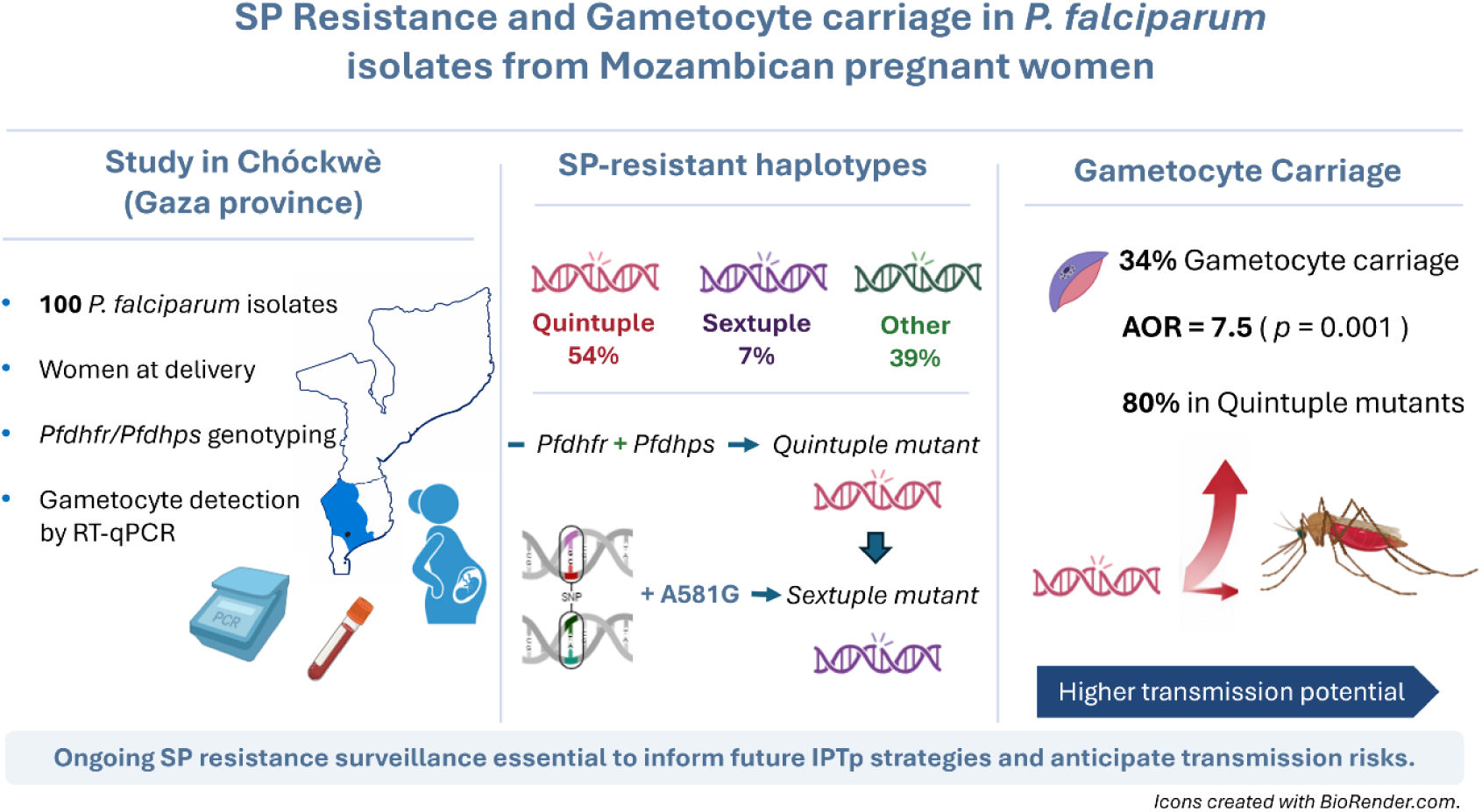

## 1. Introduction

Malaria remains a major cause of maternal and perinatal morbidity and mortality in sub-Saharan Africa (SSA). Approximately 33 million pregnant women in SSA are at risk of acquiring *Plasmodium falciparum (P. falciparum)* each year [1]. Current interventions to prevent malaria in pregnancy (MiP) and improve pregnancy outcomes include the use of long lasting insecticidal nets (LLINs), administration of intermittent preventive treatment in pregnancy with sulfadoxine-pyrimethamine (IPTp-SP) and timely treatment with effective antimalarial drugs [1,2]. IPTp-SP should be given starting from the second trimester, at monthly intervals, with a minimum of three doses continuing until delivery, at antenatal care (ANC) visits [2,3].

The effectiveness of sulfadoxine-pyrimethamine (SP) for IPTp is increasingly compromised by the emergence and spread of SP-resistant strains [4]. This resistance is primarily due to specific point mutations in the *P. falciparum* dihydrofolate reductase (*pfdhfr*) and dihydropteroate synthase (*pfdhps*) genes coding for the enzymes targeted by SP. Cumulative point mutations in the *pfdhfr* and *pfdhps* genes are associated with increased resistance to SP [5]. The quintuple mutant parasite, characterized by *pfdhfr* substitutions N51I, C59R, S108N and *pfdhps* substitutions A437G and K540E, associated with mid-level resistance, has been linked to a higher risk of SP treatment failure in malaria-infected children and a reduced prophylactic period in pregnant women [4,6–9], although protection against low birth weight (LBW) is sustained [7,10]. The emergence of the *pfdhps* A581G mutation on a quintuple mutant background, forming the so-called sextuple mutant associated with high-level resistance [11,12], has led to a reduced effectiveness of IPTp-SP in preventing malaria infections [13]. IPTp-SP remains effective at reducing LBW and maternal anemia, even in areas with high SP resistance (although the protective effect is less than in areas with lower levels of resistance), possibly through non-malaria effects on fetal growth [14]. In contrast, other studies did not find a statistically significant association between A581G mutations and reduced IPTp-SP efficacy [15,16]. Additional mutations, such as *pfdhfr* I164L and *pfdhps* S436F and A613S are associated with increased SP resistance [12,17,18].

In Mozambique, IPTp-SP was implemented in 2006 [19] and the national guidelines were updated in 2014 to implement equal or more than 3 SP-doses during pregnancy as recommended by WHO. The prevalence of the quintuple mutant haplotype in *P. falciparum*-infected individuals increased significantly over time, from 80% in 2015 to 89% in 2018, reaching 95% in Maputo (southern Mozambique) in 2018 [20]. A recent genomic surveillance study conducted in 2021-2022 reported similarly high prevalences nationwide, ranging from 87.8% to 92.7% [21]. In Gaza province, the quintuple mutant increased from 56% in 2006 to 76% in 2010 [22], and further to 89.5% in 2018 [20,23]. In contrast, the prevalences of *pfdhps* A581G and A613S, and *pfdhfr* I164L mutations remained low, below 1% nationwide in 2018 [20], and below 2% in 2021-2022 [21]. Among pregnant women receiving IPTp-SP in Maputo Province (Manhiça district) between 2016 and 2019, 94% carried the quintuple mutant haplotype. However, no *pfdhps* A581G mutation, indicative of the sextuple mutant associated with reduced SP efficacy, was observed [20,23].

SP has limited gametocytocidal activity, particularly against mature gametocytes [24–26]. Several studies have reported increased microscopic and submicroscopic gametocyte carriage following SP-administration, often at densities sufficient to infect *Anopheles* mosquitoes [27–30]. It has also been shown that SP can influence gametocyte sex ratio [31–35] in favor of more male gametocytes, which may have implications in increasing infectivity to mosquitos [33,35,36]. Therefore, even when IPTp-SP can effectively clear asymptomatic infections in pregnant women, it may simultaneously increase gametocyte carriage, alter sex ratios, and thus contribute to the human infectious reservoir [35,37]. In this context, the potential of IPTp-SP to select resistant strains through gametocyte-mediated transmission needs consideration, particularly in areas with high resistance prevalence [8,13,27,38].

In this study, we assessed the frequency of *pfdhfr/pfdhps* mutations in *P. falciparum* parasites collected from pregnant women at delivery in the rural Chókwè district, Gaza province, Mozambique, between 2014 and 2015. We analyzed associations between mutant haplotypes and parasitological and pregnancy outcomes as well as the impact of IPTp-SP on gametocyte carriage and densities.

## 2. Materials and Methods

### 2.1. Study site, population and samples

In this study, we analyzed 100 samples from pregnant women who had *P. falciparum* infection at the time of delivery and participated in a descriptive observational study published elsewhere [39]. In brief, the original study enrolled 914 pregnant women at delivery in Chókwè district, Southern Mozambique, as part of a previous hospital-based survey conducted between June 2014 and June 2015. Chókwè is endemic for *P. falciparum* and characterized by perennial malaria transmission. Detailed descriptions of the study site and population have been described before [39]. Immediately after delivery, a 3 ml venous blood sample was collected from each woman into EDTA-containing tubes. From this, 200 µl was transferred into an EDTA microtainer, and 100 µl into RNAprotect, for subsequent DNA and RNA extraction, respectively.

### 2.2. DNA extraction and *P. falciparum* diagnosis

Molecular detection of *P. falciparum* infections was performed by qPCR [40]. Briefly, DNA was extracted, according to the manufacturer’s instructions, from 200μl of erythrocyte pellet using QIAamp 96 DNA blood kit (Qiagen, Germany) and eluted in 200μl of water. Five microliters of DNA were used for qPCR analysis targeting *P. falciparum var* gene acidic terminal sequence (*var*ATS, ∼ 59 copies per genome) as previously described [40]. Parasite densities were obtained by interpolating cycle thresholds (Ct) from a standard curve of infected erythrocytes diluted in whole blood (from 100,000 to 0.01 parasites/μl of blood). Samples with Ct values ≤ 38.5 Ct were considered positive. The limit of detection was 0.04 parasites/μl of blood. Placental infections were detected in placental tissues by histological examination as described elsewhere [39].

### 2.3. Genotyping *pfdhfr* and *pfdhps* genes

To genotype mutations at the *pfdhfr* loci (N51I, C59R, S108N, and I164L) and the *pfdhps* loci (S436F, A437G, K540E, and A581G), we performed PCR-restriction fragment length polymorphism (PCR-RFLP) on a Biometra T professional gradient Thermocycler (Thistle Scientific Ltd, UK) using primers and nested-PCR protocols described previously [41,42]. Primer pairs and restriction enzymes used for *pfdhfr* and *pfdhps* polymorphism detection are described in Table S1. This method was used due to its high sensitivity to detect mixed infections, observed at high proportion in regions with moderate-to-high malaria transmission. Restriction enzyme digestions were performed on 5 µl of PCR product in a final volume of 15 μl according to the manufacturer’s instructions (New England Biolabs, Beverly, MA). Double digestions were carried out for specific codons: at codon 581 using BslI and BstUI, and at codon 164 using DraI, with reactions incubated at the temperatures and durations recommended by the manufacturer (Supplementary materials). Plasmid controls for PCR-RFLP obtained from MR4-BEI resources (https://www.beiresources.org/MR4Home.aspx) were used as wild-type and mutant controls for *pfdhfr* and *pfdhps* polymorphisms (Supplementary materials). Specific plasmids included were FR-3D7 (MRA199), FR-V1/S (MRA195), PS-FCR (MRA192), PS-Dd2 (MRA193), PS-Mali (MRA191), and PS-Peru (MRA190), as detailed in the supplementary materials. We defined mixed alleles as mutant.

### 2.4. Multiplicity of infection

To determine the multiplicity of infection (MOI), *P. falciparum* merozoite protein 1 and 2 (*pfmsp1* and *pfmsp2*) were amplified by PCR and analyzed by capillary electrophoresis (Genoscreen, Lille, France) [43,44]. MOI was defined as the highest frequency of *pfmsp1* or *pfmsp2* alleles in a single sample.

### 2.5. RNA extraction and quantification of gametocytes by *Pfs25* RT-qPCR and light microscopy

RNA extraction of *P. falciparum var*ATS positive samples was done using the RNeasy Plus 96 Kit (Qiagen, Germany) from 100μl of blood collected into 500μl of RNAprotect stabilizer reagent (Qiagen) using the manufacturer’s instructions. Extracted RNA was eluted in a final volume of 90μl of RNase-free water (Qiagen). RNA extractions were treated with on-column DNase I treatment (Qiagen) to remove DNA contaminants, according to the manufacturer’s instructions. *Pfs25* female gametocyte-specific transcripts were detected using a one-step reverse transcription qPCR (RT-qPCR) [45,46]. Details are further described in the supplementary materials. Briefly, *P. falciparum* gametocyte densities were quantified using a 7-point standard curve ranging from 10^5^ to 0.1 gametocytes/μl generated from cultured 3D7 *P. falciparum* stage V gametocytes, prepared as described elsewhere [46]. The limit of detection (LOD) of this assay was 0.1 gametocytes/µl of blood. Samples with Ct values higher than the last standard curve point, were considered to contain < 0.1 gametocytes/µl, *i.e.,* LOD. Mature *P. falciparum* gametocytes were detected in peripheral blood smears by light microscopy (LM) [41].

### 2.6. Data analysis

Data were analyzed using STATA version 14.2 (Stata Corp, College Station, TX, USA) and RStudio software (version 2025.09.1; ©2009-2025 Posit Software, PBC; RRID: SCR_000432). A significance level of *p* ≤ 0.05 was assigned to all analyses. Parity was categorized as primigravidae (first pregnancy) and multigravidae (two or more pregnancies). Low birth weight was defined as birth weight at delivery less than 2500 g. Age was categorized as < 20 and ≥ 20 years old. Binary variables for mutant haplotypes were defined as those carrying a specific mutation or haplotype versus all the others. Haplotypes were built in monoallelic infections at *dhfr* and *dhps* loci, or when only one of the haplotype positions had multiple alleles. In infections with multiple variants at more than one loci, haplotypes were estimated as “mutant haplotype”. Samples were grouped by “mutant haplotype”: quintuple mutant (triple *pfdhfr* + double *pfdhps*), sextuple mutant (quintuple + A581G), and “other” haplotypes (including triple *pfdhfr*, wild-type, and others described in Table S2).

Frequencies of point mutations and infection haplotypes were determined both overall and stratified by IPTp-SP uptake (< 3 vs. ≥ 3 doses). Associations between risk factors and mutation carriage were assessed using univariate and multivariate regression analyses. Similar models were used to evaluate associations between mutant haplotypes and adverse pregnancy outcomes, among other risk factors. Categorical variables were compared using the χ^2^ test or Fisher’s exact test, as appropriate. Continuous variables were compared using the Student’s t-test and the Kruskal-Wallis test, as appropriate.

Gametocyte density data (gametocytes/µl of blood) were strongly right-skewed and deviated from normality. Gametocyte densities were log10-transformed prior to regression analyses to reduce skewness and improve linearity, however, the transformed data remained non-normally distributed. Raw gametocyte densities are reported for descriptive and comparative purposes. Differences in gametocyte carriage across mutant haplotype groups were assessed using Fisher’s exact test, and differences in gametocyte densities across mutant haplotype and IPTp-SP uptake groups were assessed using the Kruskal-Wallis test. Frequencies of gametocyte carriers were also compared among IPTp-SP uptake groups (none, one, two vs. ≥ 3 doses) using the χ^2^ test. Median [IQR] gametocyte densities were visualized using dot plots.

Univariate and multivariate logistic regression models were used to examine risk factors associated with mutant (SP-resistant) haplotypes and gametocyte carriage. Linear regression models were applied to assess associations with gametocyte densities and the effect of mutant haplotypes on pregnancy outcomes. Model assumptions for linear regression were evaluated by visual inspection of residual diagnostic plots. Effect estimates from linear models are presented on the log scale, corresponding to multiplicative differences in gametocyte density.

## 3. Results

### 3.1. Characteristics of the study participants

The demographic and parasitological characteristics of the 100 pregnant women with a *P. falciparum* infection at delivery included in this study are described in Table 1. Seventy-three percent (73/100) of women had a submicroscopic *P. falciparum* infection (positive by qPCR and negative by LM) at delivery, and 54% (54/100) of women received ≥ 3 IPTp-SP doses. Submicroscopic infections at delivery were present in 75.6% (65/86) of women that received at least one dose of IPTp-SP and 57.1% (8/14) among those that did not receive IPTp (*p* = 0.15). MOI was successfully determined in 95 samples with median [n[IQR] MOI of 3.00[2.0-4.0] clones/sample. Polyclonal infections were found in 83% of cases (83/95). There were 5 samples that could not be amplified to assess MOI (Table 1).

**Table 1.**
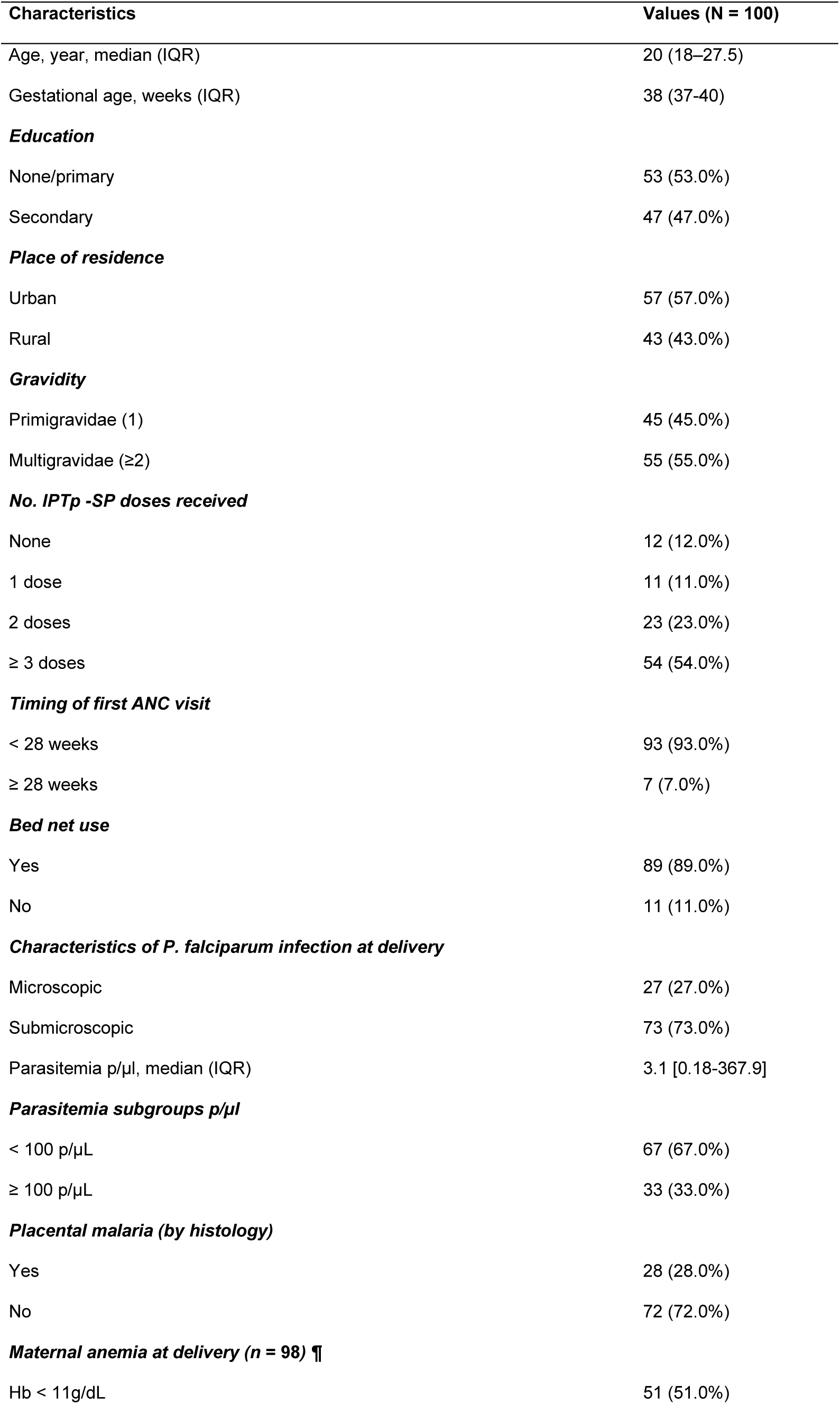
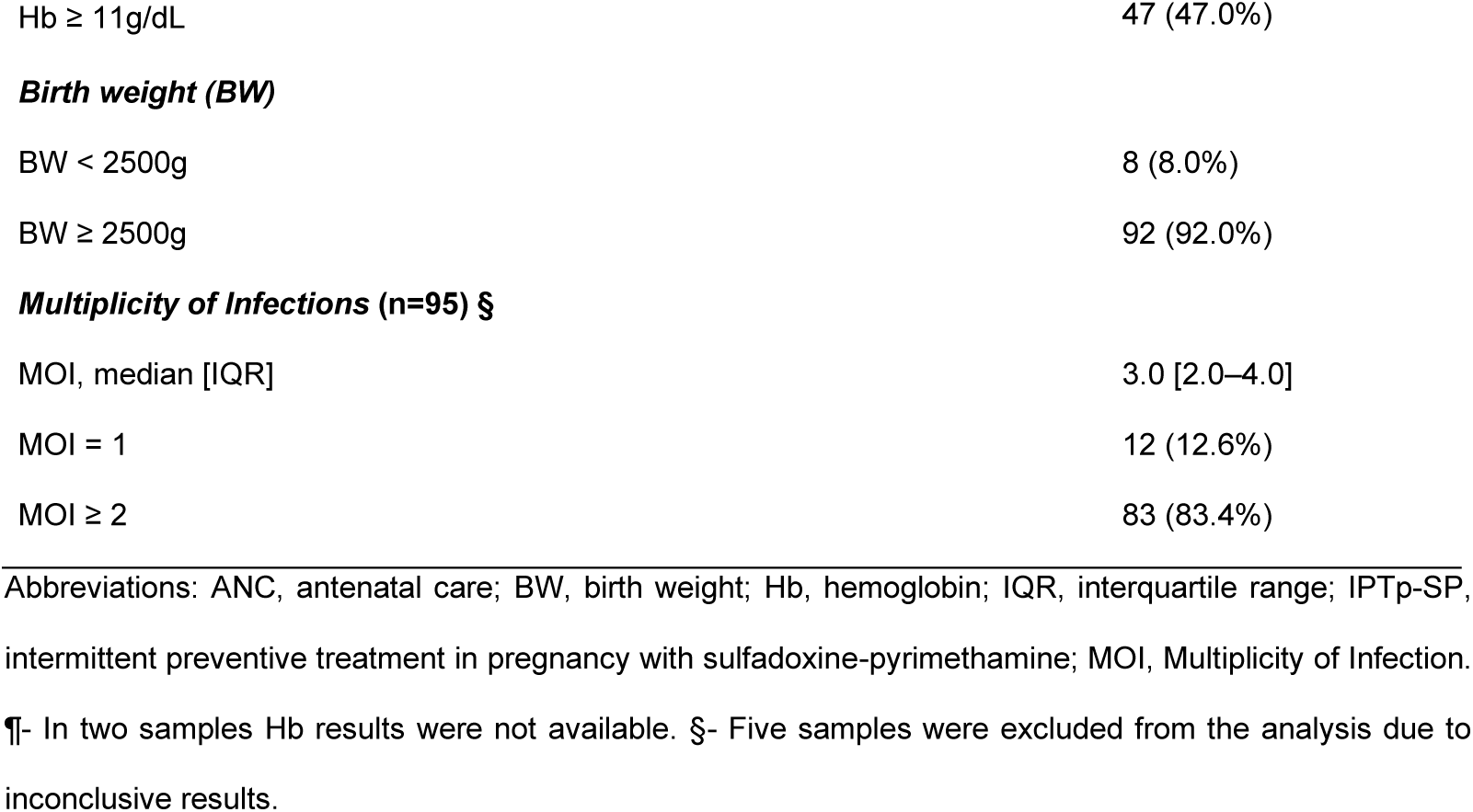
Demographic and parasitological characteristics of the study population.

| Characteristics | Values (N = 100) |
| --- | --- |
| Age, year, median (IQR) | 20 (18–27.5) |
| Gestational age, weeks (IQR) | 38 (37–40) |
| **Education** |  |
| None/primary | 53 (53.0%) |
| Secondary | 47 (47.0%) |
| **Place of residence** |  |
| Urban | 57 (57.0%) |
| Rural | 43 (43.0%) |
| **Gravidity** |  |
| Primigravidae (1) | 45 (45.0%) |
| Multigravidae (≥2) | 55 (55.0%) |
| **No. IPTp -SP doses received** |  |
| None | 12 (12.0%) |
| 1 dose | 11 (11.0%) |
| 2 doses | 23 (23.0%) |
| ≥ 3 doses | 54 (54.0%) |
| **Timing of first ANC visit** |  |
| < 28 weeks | 93 (93.0%) |
| ≥ 28 weeks | 7 (7.0%) |
| **Bed net use** |  |
| Yes | 89 (89.0%) |
| No | 11 (11.0%) |
| **Characteristics of *P. falciparum* infection at delivery** |  |
| Microscopic | 27 (27.0%) |
| Submicroscopic | 73 (73.0%) |
| Parasitemia p/μl, median (IQR) | 3.1 [0.18–367.9] |
| **Parasitemia subgroups p/μl** |  |
| < 100 p/μL | 67 (67.0%) |
| ≥ 100 p/μL | 33 (33.0%) |
| **Placental malaria (by histology)** |  |
| Yes | 28 (28.0%) |
| No | 72 (72.0%) |
| **Maternal anemia at delivery (n = 98) ¶** |  |
| Hb < 11g/dL | 51 (51.0%) |
| Hb ≥ 11g/dL | 47 (47.0%) |
| **Birth weight (BW)** |  |
| BW < 2500g | 8 (8.0%) |
| BW ≥ 2500g | 92 (92.0%) |
| **Multiplicity of Infections (n=95) §** |  |
| MOI, median [IQR] | 3.0 [2.0–4.0] |
| MOI = 1 | 12 (12.6%) |
| MOI ≥ 2 | 83 (83.4%) |

Abbreviations: ANC, antenatal care; BW, birth weight; Hb, hemoglobin; IQR, interquartile range; IPTp-SP,
intermittent preventive treatment in pregnancy with sulfadoxine-pyrimethamine; MOI, Multiplicity of Infection.
¶- In two samples Hb results were not available. §- Five samples were excluded from the analysis due to
inconclusive results.

### 3.2. Frequency of P. falciparum pfdhfr-pfdhps alleles

Genotyping results are summarized in Table 2, Table 3 and Table S2.

**Table 2.** Prevalence of P. falciparum dhfr and dhps mutant alleles per isolate.

| Gene | Mutation | N (Mutation prevalence) |
| --- | --- | --- |
| *dhfr* | N51I | 89 (89%) |
|  | C59R | 86 (86%) |
|  | S108N | 90 (90%) |
|  | I164L | 2 (2%) |
| *dhps* | A437F | 1 (1%) |
|  | A437G | 74 (74%) |
|  | K540E | 79 (79%) |
|  | A581G | 14 (14%) |
|  | A613S | 10 (10%) |

Abbreviations: *dhfr*, dihydrofolate reductase gene; *dhps*, dihydropteroate synthetase gene.

**Table 3.** Prevalence of P. falciparum dhfr and dhps combined mutation haplotypes.

| Haplotype type | Mutation Alleles | N (prevalence) |
| --- | --- | --- |
| *dhfr* | — (wildtype) | 8 (8,0%) |
|  | N51I/C59R/S108N | 80 (80,0%) |
|  | N51I/C59R/S108N/I164L | 2 (2,0%) |
|  | — (others)* | 10 (10%) |
| *dhps* | — (wildtype) | 11 (11,0%) |
|  | A437G/K540E | 56 (56,0%) |
|  | A437G/K540E/A581G | 10 (10,0%) |
|  | — (others)* | 23 (23%) |
| *dhfr/dhps* | N51I/C59R/S108N + A437G/K540E (Quintuple) | 54 (54,0%) |
|  | N51I/C59R/S108N+A437G/K540E/A581G (Sextuple) | 7 (7,0%) |
|  | N51I/C59R/S108N + A437G/K540E/A163S | 3 (3,0%) |
|  | — (others)* | 36 (36%) |

\*Composition of other mutations and combinations is presented in Table S2. **Abbreviations:** *dhfr*, dihydrofolate reductase gene; *dhps*, dihydropteroate synthetase gene.

Mutations in the *pfdhfr* gene were found in 92% of isolates, with frequencies of 89% for N51I, 86% for C59R, 90% for S108N, and 2% for I164L (Table 2). Seventy-nine percent of isolates carried the *pfdhfr* triple mutant haplotype (IRN [N51I, C59R, S108N]) with two samples carrying the additional I164L mutation (Table 3). Mutations in the *pfdhps* gene were found in 91.0% of isolates: mutations A437G and K540E were detected in 74% and 79% of isolates, respectively, while the double mutation (GE [A437G, K540E]) were detected in 57% of isolates. The A581G mutation was found in 14% of samples, 71.4% (10/14) of which carried the triple mutant haplotype (GEG [A437G, K540E, and A581G]) and were polyclonal infections. Among those, in 28.6% (4/14) of isolates the MOI was not calculated as indicated in Table 1.

After combining *pfdhfr-pfdhps* haplotypes, the quintuple mutant haplotype (IRN-GE) was observed in 54% (54/100) of isolates. The sextuple mutant haplotype (IRN-GEG) was observed in 7% (7/100) isolates, while only 2% (2/100) of isolates did not have any mutation (wild-type).

### 3.3. Risk factors for carriage of quintuple and sextuple mutant parasites

We investigated risk factors associated with quintuple and sextuple mutation carriage. Results from the multivariate analysis are shown in Table 4 (the univariate analysis are shown in Table S3). Parasite density at delivery (peripheral parasite density ≥ 100 p/µl) was not associated with increased carriage of resistant parasites. However, pregnant women receiving ≥ 3 IPTp-SP doses had higher odds (AOR = 3.7, *p* < 0.001) of carrying quintuple mutant parasites (IRN-GE), while no association was observed in women carrying the sextuple mutant parasites.

**Table 4.**
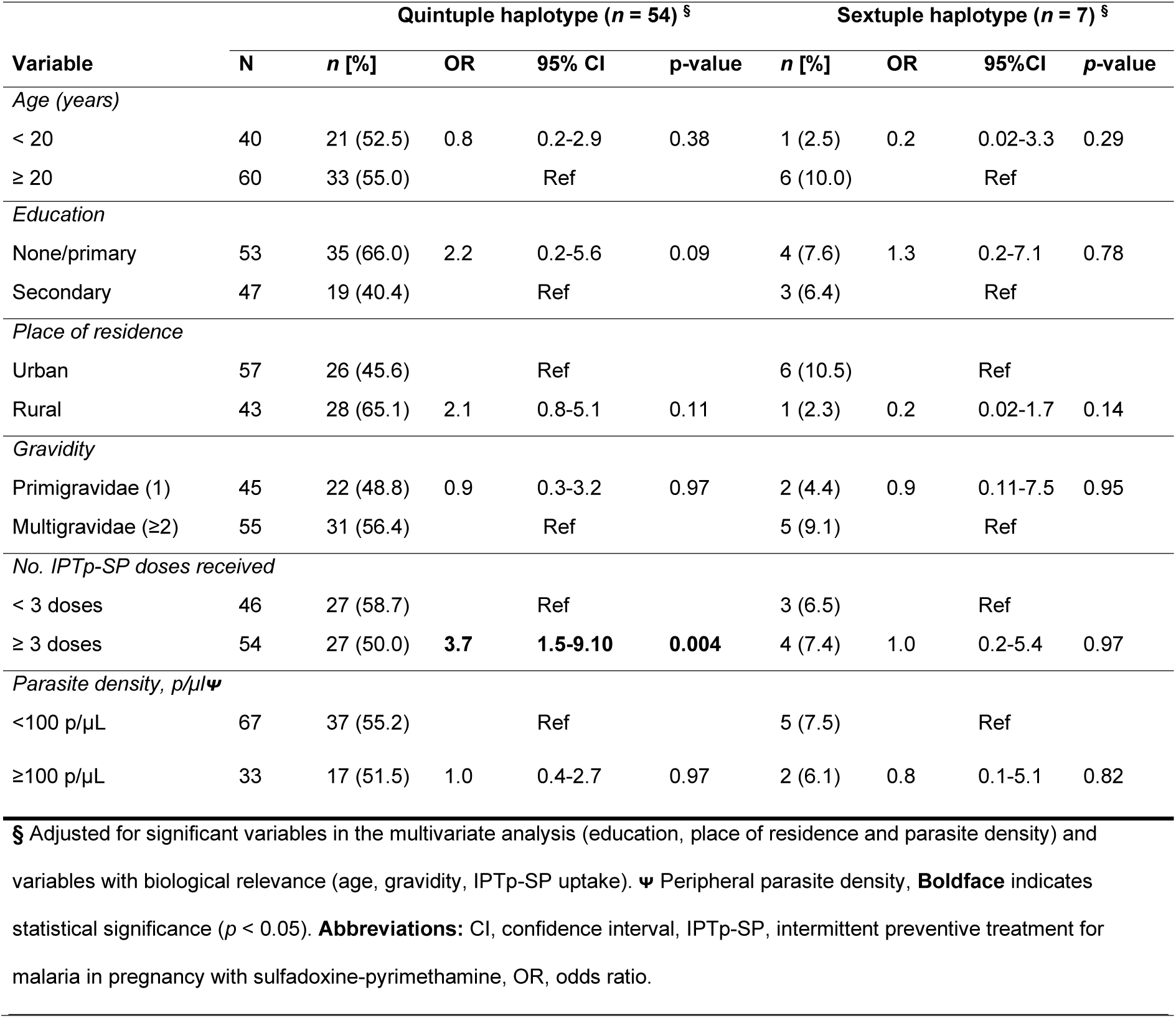
Multivariate analysis of risk factors associated with carriage of mutant haplotypes (N = 100).

| Variable | Quintuple haplotype (n = 54) § |  |  |  |  | Sextuple haplotype (n = 7) § |  |  |  |
| --- | --- | --- | --- | --- | --- | --- | --- | --- | --- |
|  | N | n [%] | OR | 95% CI | p-value | n [%] | OR | 95%CI | p-value |
| *Age (years)* |  |  |  |  |  |  |  |  |  |
| < 20 | 40 | 21 (52.5) | 0.8 | 0.2-2.9 | 0.38 | 1 (2.5) | 0.2 | 0.02-3.3 | 0.29 |
| ≥ 20 | 60 | 33 (55.0) |  | Ref |  | 6 (10.0) |  | Ref |  |
| *Education* |  |  |  |  |  |  |  |  |  |
| None/primary | 53 | 35 (66.0) | 2.2 | 0.2-5.6 | 0.09 | 4 (7.6) | 1.3 | 0.2-7.1 | 0.78 |
| Secondary | 47 | 19 (40.4) |  | Ref |  | 3 (6.4) |  | Ref |  |
| *Place of residence* |  |  |  |  |  |  |  |  |  |
| Urban | 57 | 26 (45.6) |  | Ref |  | 6 (10.5) |  | Ref |  |
| Rural | 43 | 28 (65.1) | 2.1 | 0.8-5.1 | 0.11 | 1 (2.3) | 0.2 | 0.02-1.7 | 0.14 |
| *Gravidity* |  |  |  |  |  |  |  |  |  |
| Primigravidae (1) | 45 | 22 (48.8) | 0.9 | 0.3-3.2 | 0.97 | 2 (4.4) | 0.9 | 0.11-7.5 | 0.95 |
| Multigravidae (≥2) | 55 | 31 (56.4) |  | Ref |  | 5 (9.1) |  | Ref |  |
| *No. IPTp-SP doses received* |  |  |  |  |  |  |  |  |  |
| < 3 doses | 46 | 27 (58.7) |  | Ref |  | 3 (6.5) |  | Ref |  |
| ≥ 3 doses | 54 | 27 (50.0) | **3.7** | **1.5-9.10** | **0.004** | 4 (7.4) | 1.0 | 0.2-5.4 | 0.97 |
| *Parasite density, p/μL* |  |  |  |  |  |  |  |  |  |
| <100 p/μL | 67 | 37 (55.2) |  | Ref |  | 5 (7.5) |  | Ref |  |
| ≥100 p/μL | 33 | 17 (51.5) | 1.0 | 0.4-2.7 | 0.97 | 2 (6.1) | 0.8 | 0.1-5.1 | 0.82 |

§ Adjusted for significant variables in the multivariate analysis (education, place of residence and parasite density) and variables with biological relevance (age, gravidity, IPTp-SP uptake). $\Psi$ Peripheral parasite density, **Boldface** indicates statistical significance ( $p < 0.05$ ). **Abbreviations:** CI, confidence interval, IPTp-SP, intermittent preventive treatment for malaria in pregnancy with sulfadoxine-pyrimethamine, OR, odds ratio.

We also investigated whether the odds of adverse pregnancy outcomes, *i.e.* LBW, placental malaria and pre-term delivery was higher in women carrying quintuple and sextuple mutant parasites at delivery. Results from the multivariate analysis did not show significant associations between the carriage of resistant parasites and adverse outcomes (Table 5; the univariate analysis is presented in Table S4).

**Table 5.**
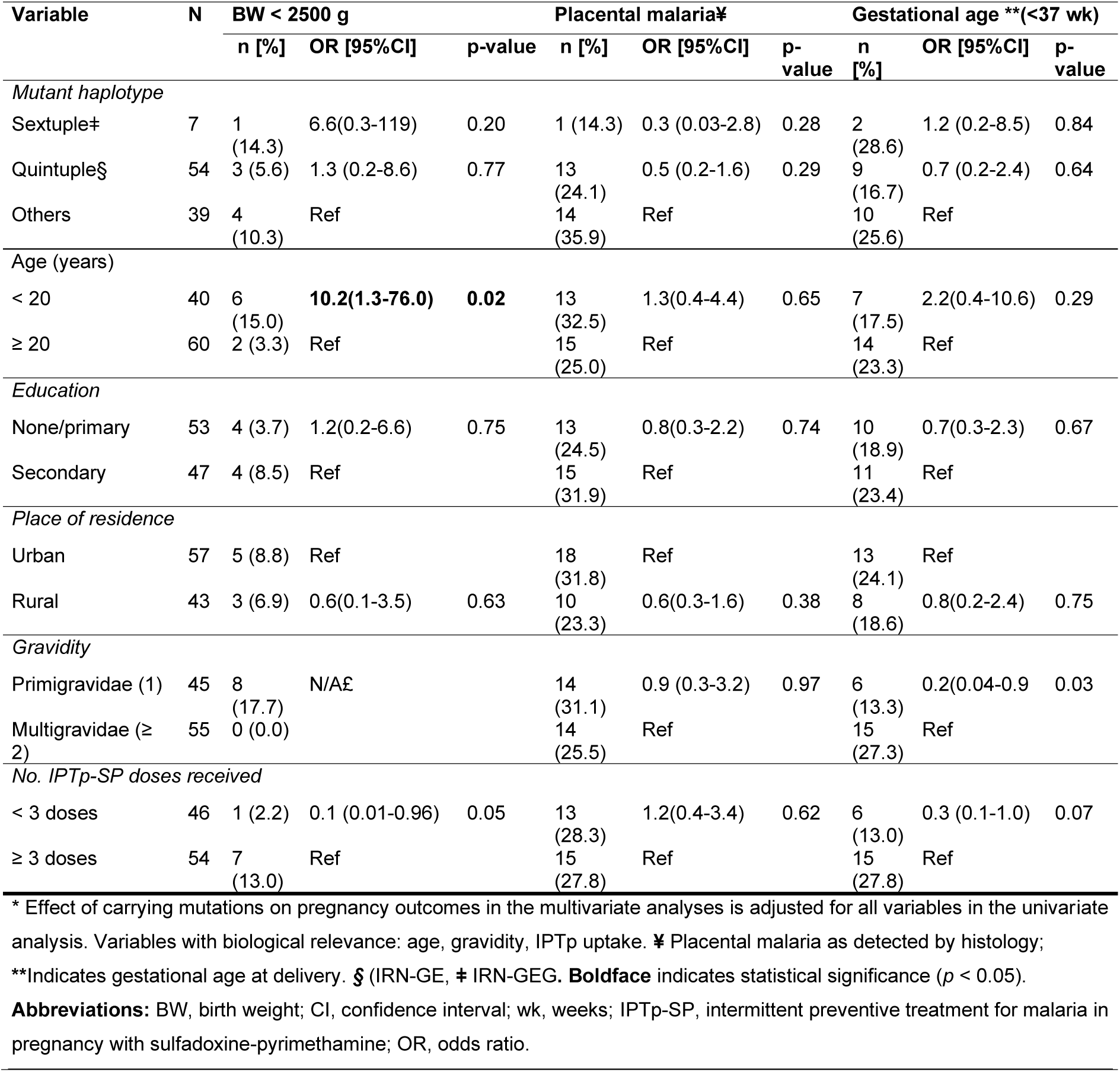
Effect of mutant haplotypes on adverse pregnancy outcomes in Chókwè district (N = 100) * (Multivariate analysis).

There was no significant difference in the proportion of monoclonal (MOI = 1) and polyclonal (MOI ≥ 2) infections between participants carrying parasites with no mutations and those carrying parasites with one or more mutations (*p* = 0.63). The distribution of *pfmsp1* and *pfmsp2* allelic families did not significantly differ among study participants carrying parasites with no mutations (wild type) compared to those with one or more mutations.

The distribution of *pfmsp1* and *pfmsp2* allelic families did not significantly differ among study participants carrying parasites with no mutations compared to those with one or more mutations. The median MOI across mutant haplotype groups was 4.5 [2.5-5.5] in “wildtype” parasites, 3 [2.0-3.0] in study participants carrying “quintuple (*dhfr/dhps*),” and 3 [2.0-3.0] “sextuple (*dhfr/dhps*)” parasites, and 3.5 [2.5-5.0] in participants carrying in parasites with “other haplotypes” (Supplementary file Table S5).

### 3.4. IPTp-SP and gametocyte carriage

*Pfs25* transcripts were detected in 34/100 women at delivery using one-step RT-qPCR, resulting in an overall gametocyte carriage prevalence of 34% in the study population. In contrast, only 2/100 women were positive for gametocytes by LM. Most gametocyte carriers harbored parasites with quintuple mutations (26/34, 80%) (Table 6).

**Table 6.** Gametocyte carriage and densities by mutant haplotype.

|  | Mutant haplotype |  |  |  | *p*-value |
| --- | --- | --- | --- | --- | --- |
|  | Total<br>N = 100 | Other°<br>n = 39 | Quintuple<br>n = 54 | Sextuple<br>n = 7 |  |
| Gametocyte carrier n (n/N %) | 34 (34.0) | 6 (15.4) | 27 (50.0) | 1 (14.3) | **0.0011** |
| No gametocyte carriers with densities<br>< LOD | 19 | 3 | 15 | 1 |  |
| No gametocyte carriers with densities<br>> LOD | 15 | 3 | 12 | 0 |  |
| Median gametocyte density (n = 15) | 1.76 | 0.57 | 1.81 | NA | 0.31 |
| Interquartile range [IQR] | [0.62-3.19] | [0.36-1.75] | [0.76-3.92] | [NA-NA] |  |

Gametocyte carriage was more prevalent in infections with quintuple mutant parasites compared to sextuple and other haplotypes (Fisher’s exact test, *p* = 0.0011; Table 6). Median [IQR] gametocyte densities were low across all haplotype groups (1.76 [0.62-3.19]) and did .not differ significantly between haplotypes (Kruskal-Wallis test, *p* = 0.31; Figure 1, Table 6). Similarly, gametocyte carriage and densities did not differ significantly across IPTp-SP dose groups (χ^2^ test, *p* = 0.40; Kruskal-Wallis test, *p* = 0.39, respectively) (Table S6)

**Figure 1.**
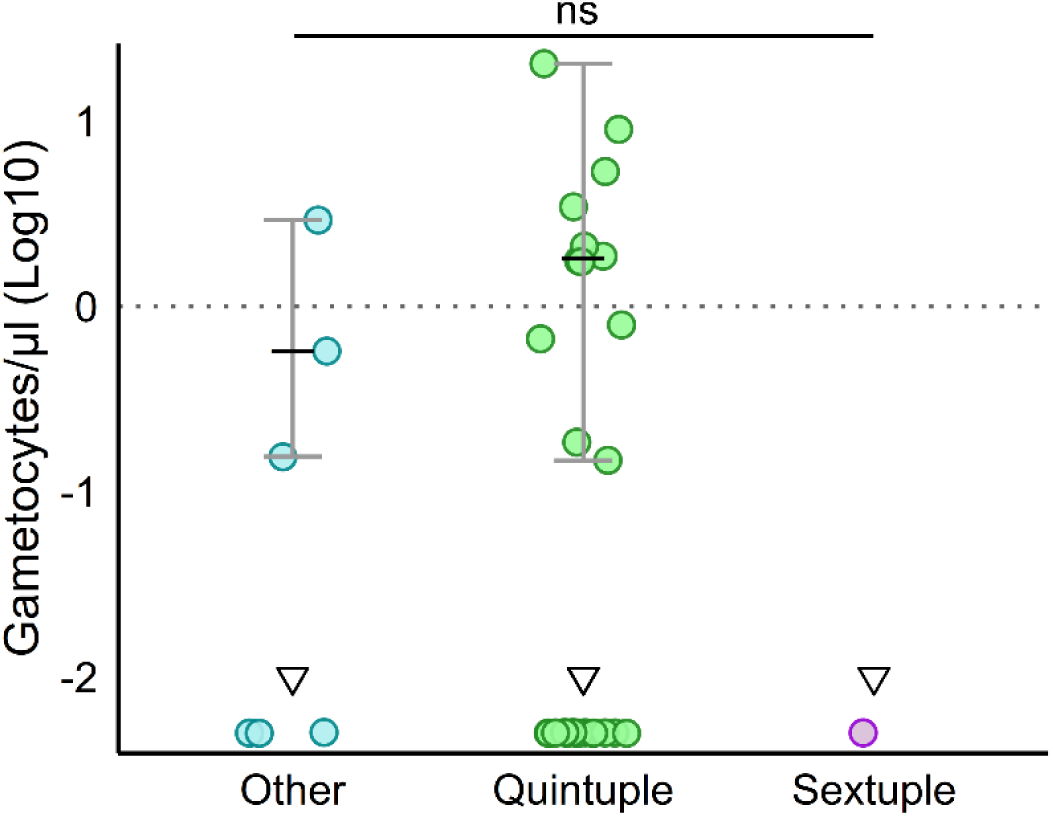
Gametocyte densities by mutant haplotype. Individual-level log10-transformed gametocyte densities (gametocytes/µl of blood) are visualized (Y-axis), amongst 34 gametocyte carriers, with each dot representing one individual sample. Medians (black and bold horizontal lines) and interquartile ranges (grey error bars) were overlaid per group in samples above LOD (≥ 0.01 gametocytes/µl). Samples below the LOD (< 0.01 gametocytes/µl) are included and indicated with dots with on top a downward-facing triangle by haplotype. Mutant haplotypes are ordered as “Other” haplotypes i.e., “Triple pfdhfr” and wild-type among others described in Table S2 (n = 6, 3 above and 3 below LOD), “Quintuple (dhfr/dhps)” (n = 27, 12 above and 15 below LOD), and “Sextuple (dhfr/dhps)” (n = 1 below LOD). Data points represent individual samples. Non-significant p-value (p = 0.31) is indicated with a large horizontal bold line (Kruskal-Wallis test). Abbreviations: dhfr, dihydrofolate reductase; dhps, dihydropteroate synthase; IQR, interquartile range; LOD, limit of detection by RT-qPCR; RT-qPCR, reverse transcription quantitative polymerase chain reaction; SP, sulfadoxine-pyrimethamine.

To investigate factors associated with gametocyte carriage, we performed a multivariate logistic regression analysis (Table 7). Mutant haplotype was the only significant predictor: women with infections with quintuple mutant parasites had 7.5-fold higher odds of carrying gametocytes (AOR = 7.5, 95% CI 2.5-27.3, *p* = 0.001). Age, place of residence, gravidity, IPTp-SP dose, parasite density, and placental malaria were not significantly associated with gametocyte carriage in the adjusted models (Table 7).

**Table 7.**
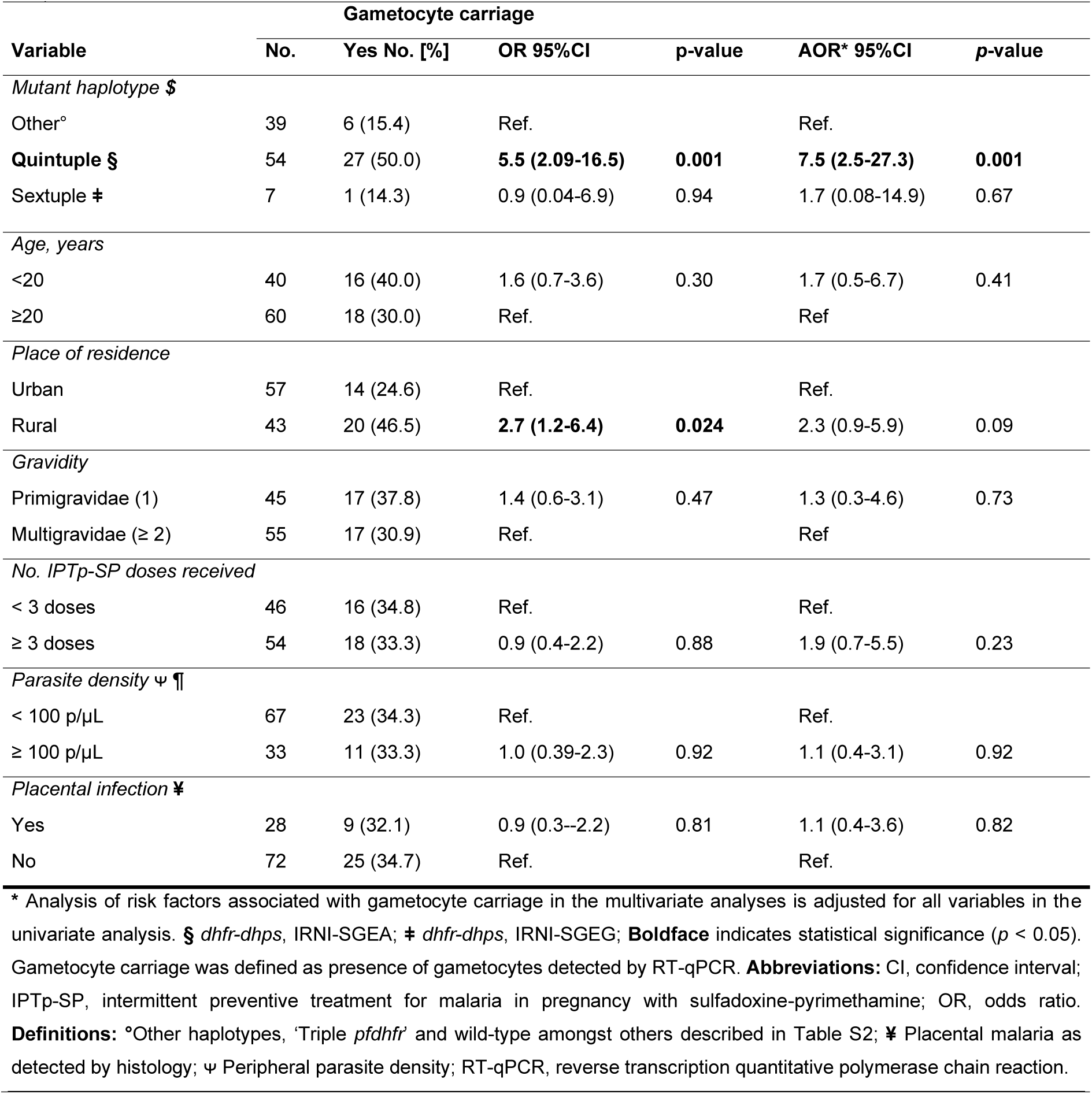
Univariate and multivariate logistic regression analysis of risk factors associated with gametocyte carriage* (N = 100).

We further investigated factors associated with gametocyte density, modeled as a continuous variable in univariate linear regression. However, none of the variables included in the analysis were significantly associated with gametocyte density (Table 8), indicating that age, place of residence, gravidity, number of IPTp-SP doses, parasite density, placental infection, or the presence of SP resistance markers may not be drivers of increased gametocyte densities among pregnant women in our study population.

**Table 8.** Univariate and multivariate linear regression analysis of risk factors associated with increased gametocyte densities* (N = 100).

| Variable | Gametocyte density (log10, gametocytes/μl) |  |  |  |
| --- | --- | --- | --- | --- |
|  | n | n/N [%] | Mean (SD) | Coefficient (95%CI, p-value) (Univariate) |
| *Mutant haplotype §* |  |  |  |  |
| Other° | 3 | 20.0 | 0.3 (0.2) | Ref. |
| Quintuple § | 12 | 80.0 | 0.5 (0.1) | 0.23 (-0.27 to 0.74, p = 0.34) |
| Sextuple ‡ | 0 | 00.0 | NA | NA |
| *Age, years* |  |  |  |  |
| < 20 | 8 | 50.0 | 0.6 (0.4) | 0.33 (-0.04 to 0.70, p = 0.08) |
| ≥ 20 | 7 | 50.0 | 0.3 (0.2) | Ref. |
| *Place of residence* |  |  |  |  |
| Urban | 3 | 20.0 | 0.6 (0.4) | Ref. |
| Rural | 12 | 80.0 | 0.4 (0.3) | -0.21 (-0.62 to 0.20, p = 0.30) |
| *Gravidity* |  |  |  |  |
| Primigravidae (1) | 9 | 60.0 | 0.6 (0.4) | 0.22 (-0.19 to 0.62, p = 0.27) |
| Multigravidae (≥2) | 6 | 40.0 | 0.4 (0.3) | Ref. |
| *No. IPTp doses received* |  |  |  |  |
| < 3 doses | 9 | 60.0 | 0.6 (0.4) | Ref. |
| ≥ 3 doses | 6 | 40.0 | 0.4 (0.3) | -0.24 (-0.65 to 0.17, p = 0.22) |
| *Parasite density ψ ¶* |  |  |  |  |
| < 100 p/μL | 6 | 40.0 | 0.5 (0.4) | Ref. |
| ≥ 100 p/μL | 9 | 60.0 | 0.2 (0.1) | -0.37 (-0.85 to 0.11, p = 0.12) |
| *Placental infection ¥* |  |  |  |  |
| Yes | 3 | 20.0 | 0.3 (0.2) | -0.21 (-0.62 to 0.20, p = 0.28) |
| No | 12 | 80.0 | 0.6 (0.4) | Ref. |

\*Analysis of risk factors associated with gametocyte densities in the multivariate analyses is adjusted for all variables in the univariate analysis, only samples with gametocyte densities above the limit of detection (LOD) are included (> 0.1 gametocytes/μl). Gametocyte densities were measured by RT-qPCR. **Abbreviations:** CI, confidence interval; IPTp-SP, intermittent preventive treatment for malaria in pregnancy with sulfadoxine-pyrimethamine; OR, odds ratio. **Definitions:** § *dhfr-dhps*, IRNI-SGEA; ‡ *dhfr-dhps*, IRNI-SGEG; NA: not able to quantify n= 0 or 1; °Other haplotypes, 'Triple *pf dhfr*' and wild-type amongst others described in Table S2; ¥ Placental malaria as detected by histology; ψ Peripheral parasite density; RT-qPCR, reverse transcription quantitative polymerase chain reaction.

## 4. Discussion

Although SP is no longer recommended for the treatment of *P. falciparum* malaria due to widespread resistance, IPTp-SP remains the main strategy to prevent MiP across most SSA countries [47]. Even in areas with a high prevalence of *pfdhfr and pfdhps* mutations, IPTp-SP continues to reduce LBW and maternal anemia, despite its reduced efficacy in preventing infection [8]. Regular monitoring of SP molecular resistance markers therefore remains essential.

In this study, 98 of 100 *P. falciparum* samples from pregnant women carried mutations in *pfdhfr* or *pfdhps* genes. Mutations I164L and A581G were detected in 2% and 14% of samples, respectively. The quintuple mutant haplotype (*pfdhfr* N51I, C59R, S108N and *pfdhps* [A437G, K540E; IRN-GE) was prevalent in 54% of infections, consistent with earlier reports from Gaza Province in 2006 [22], while the sextuple mutant haplotype (*pfdhfr* N51I, C59R, S108N and *pfdhps* A437G, K540E, A581G, IRN-GEG) occurred in 7% of infections. Our results align with the increasing prevalence over time of the quintuple mutant haplotype reported in previous surveys in Mozambique (2006-2022), with frequencies rising from 56.2% in 2006 to >87% in 2022 [10,21,22], while the additional *pfdhps* A581G mutation remained uncommon (≤1.6%) [10,21]. The higher A581G frequency observed in our study (14%) may reflect ongoing local selection pressure under sustained IPTp-SP use. Relatively high IPTp-SP coverage reported in parts of Mozambique (approximately 47-63% coverage of ≥ 3 doses) may contribute to this elevated A581G prevalence [39,48].

The predominance of quintuple mutant haplotypes in our study is consistent with patterns across East and Southern Africa, where the combination of the *pfdhfr* triple mutant (N51I, C59R, S108N) and *pfdhps* (A437G, K540E) −defining the quintuple mutant haplotype− is highly prevalent, though frequencies vary geographically [49]. However, in a study of children in Gaza Province, the A581G mutation was not present [50], suggesting that drug exposure during pregnancy exerts distinct selection pressures. A Ghanaian study identified a correlation between A581G-carrying parasites and elevated plasma SP concentrations during delivery, though mutant frequencies did not differ between women receiving ≥ 3 versus < 3 IPTp-SP doses [7,51–55], illustrating that drug pressure influences A581G selection differently across regions.

The emergence of sextuple *pfdhfr/pfdhps* haplotype has been associated with progressive reductions in IPTp-SP’s efficacy against infection [14]. Nevertheless, the protective effect on LBW persists in several regions, potentially due to SP’s non-parasitic activity via anti-inflammatory or antibacterial effects [7,14,56,57]. Our findings showed no association between sextuple haplotypes and LBW. This is consistent with existing research suggesting that the efficacy of IPTp-SP is determined by broader, population-level resistance patterns rather than the presence of specific individual haplotypes [14]. Importantly, IPTp-SP has been shown to provide benefits in Mozambique [10], indicating that it remains effective despite the presence of SP resistance markers.

Participants infected with quintuple −but not sextuple− mutants had higher asexual parasite densities. Similar findings have been reported where K540E prevalence exceeds 90%, compromising IPTp-SP’s ability to clear infections [58]. Although our K540E frequency was slightly lower (79%), these higher densities may indicate reduced SP clearance and prolonged infection duration. Parasite density, however, also reflects host immunity and parity, so these associations should be interpreted cautiously. Persistent infections contribute both to adverse outcomes [59] and to continued transmission.

The *pfdhfr* I164L mutation, detected in 2% of isolates, has not previously been reported in Mozambique [21,23,60], but has been observed at low frequencies in Tanzania [12,61,62], Uganda [12,63,64], Ghana and Burkina Faso [65]. I164L is known to substantially increase pyrimethamine resistance when present on the *pfdhfr* triple-mutant background (N51I/C59R/S108N) [66]. Although it was not detected in combination with a fully resistant *pfdhfr/pfdhps* background in our study, its emergence may represent an early step toward higher-level SP resistance, underscoring the importance of continued molecular surveillance [14,49].

The reduced effectiveness of IPTp-SP in areas with high resistance is largely explained by the SP’s long elimination half-life, which maintains subtherapeutic concentrations that suppress sensitive parasites while allowing resistant genotypes to persist and expand [67–69]. In pregnant women, intermittent SP dosing, partial immunity, and asymptomatic parasitemia further promote survival and transmission of resistant parasites [70,71].

At delivery, all women were asymptomatic; 73% had submicroscopic infections and 34% carried gametocytes detected only by RT-qPCR. Among gametocyte carriers, 80% harbored quintuple and 2.9% sextuple haplotype. These findings suggest that while SP can lower parasite density, it may fail to act as a preventative measure against new infections or as an effective curative treatment for drug-resistant strains. Asymptomatic infected women thus represent a non-negligible human reservoir, potentially contributing to the spread of resistant parasites [55,72,73].

The main predictor of gametocyte carriage in this study, was infection with the quintuple mutant haplotype, consistent with previous reports showing that emergence of SP resistance is associated with greater gametocytemia [74], while no association was observed with IPTp-SP use [28,75,76]. We also observed higher asexual parasite densities in quintuple mutant infections, and previous studies have shown that gametocyte carriage is associated with asexual parasite density [27,77]. This suggests that pregnant women infected with resistant strains are more likely to have higher asexual parasite densities, which in turn are associated with a greater likelihood of gametocytemia [30]. However, gametocyte carriage was assessed only at delivery, limiting our ability to evaluate temporal dynamics or the cumulative effect of repeated SP use on gametocyte conversion.

Evidence indicates that even low-density gametocyte carriage can contribute to malaria transmission, reflecting a complex relationship between gametocyte density and mosquito infection [34]. Lower gametocyte densities following SP treatment may partly reflect mobilization of sequestered bone marrow gametocytes into peripheral circulation [78]. Unlike artemisinin derivatives, SP has not been shown to induce gametocyte conversion *in vitro* [32,79]. However, some studies have reported a male-biased gametocyte sex ratio following SP treatment [35]. This pattern likely reflects preferential survival of male gametocytes after drug exposure rather than direct induction by SP. Since gametocyte sex ratio critically determines human-to-mosquito infectivity [38], longitudinal studies combining repeated sampling with gametocyte quantification and sex ratio analysis after SP exposure, such as those conducted in Nigeria [31] and South Africa [74], are essential. Comparable studies in Burkina Faso have linked gametocyte dynamics to mosquito infectivity independently of SP exposure [80]. Such studies are needed in Mozambique to elucidate how SP exposure shapes gametocyte biology and transmission potential [33].

In conclusion, the emergence of highly resistant malaria strains in Mozambique indicates that the standard treatment (SP) may be failing to clear infections and diminishing benefit from additional doses as resistance increases across African regions. The considerable burden of submicroscopic gametocyte carriers infected with resistant parasites suggests that pregnant women may act as a significant reservoir for transmission of resistant malaria back into the community [14]. The high rate of genetic mutations found in parasites at delivery time suggests resistance to SP is a growing threat for the current prevention strategy for protecting mothers or their babies from malaria.

## S1 METHODS

*Pfdhfr* and *pfdhps* genotyping – Restriction fragment length polymorphism.

Reverse transcription quantitative PCR for detection of Pfs25 gametocyte-specific transcripts.

## S2 TABLES

**Table S1:** Sequence of primer pairs and restriction enzymes used for *pfdhfr* and *pfdhps* polymorphism detection, size of amplicons and covered amino acids, and assessment of *Pfs*25 gametocyte-specific transcripts by RT-qPCR.

**Table S2:** Prevalence of *P. falciparum dhfr* and *dhps* haplotype combinations found at a low frequency.

**Table S3:** Univariate analysis of risk factors associated with carriage of quintuple and sextuple mutant haplotypes (*n* = 100).

**Table S4:** Effect of mutant haplotypes and other risk factors on adverse pregnancy outcomes in Chókwè district (*n* = 100) (Univariate analysis)

**Table S5:** Parasite variability, shown as Multiplicity of infection (MOI) in different mutation haplotypes.

**Table S6:** Frequency of gametocyte carriage and gametocyte densities by IPTp-SP uptake group.

**References** [42,45,46,81] are cited in the supplementary materials.

## Author contributions

Conceptualization, P.A., Y.D-EB., E.R-V and A.R-U.; Methodology, P.A., Y.D-EB. and A.R-U.; Validation, P.A., Y.D-EB., and E.R-V.; Formal Analysis, P.A. and Y.D-EB.; Investigation, P.A. Y.D-EB., J.H-K., P.G., E.R-V and D.C.; Resources, P.A., Y.D-EB., A.R-U., J.H-K., and S.M-E.; Data Curation, P.A. and Y.D-EB.; Writing – Original Draft Preparation, P.A., Y.D-EB. and A.R-U.; Writing – Review & Editing, P.A., Y.D-EB., A.R-U., E.R-V., J.H-K., L.K., and S.M-E.; Visualization, P.A. and Y.D-EB.; Supervision, A.R-U., L.K., and S.M-E.; Project Administration, A.R-U., L.K., and S.M-E.; Funding Acquisition, P.A., A.R-U., J.H-K., and Y.D-EB.. All authors read and approved of the final manuscript.

## Institutional Review Board Statement

The study was conducted in accordance with the Declaration of Helsinki, and the protocol was approved by the Ethics Committee of National Mozambican Ethics review committee (CNBS)(protocol code IRB 00002657 on the 23^th^ November 2013), the institutional review boards (IRBs) of the Institute of Tropical Medicine (protocol code IRB AB/ac/059 on the 25^th^ March 2014), and the University of Antwerp (protocol code IRB B300201421228 on the 26^th^ May 2014).

## Informed Consent Statement

Informed consent was obtained from all subjects involved in the study or their legal representatives.

## Data Availability

The original data presented in the study are openly available in Zenodo at DOI: https://doi.org/10.5281/zenodo.18788858 (anonymized dataset). Due to the sensitive nature of the data, the authors are unable to share demographic and health data directly. Requests for data access should be requested to Institute of Tropical Medicine’s (ITM’s) data access contact point. All requests will be reviewed by ITM’s Data Access Committee, which will also manage approved data sharing.

## Acknowledgments

The authors are very grateful to all the pregnant women who participated in this study, to all the nurses from the participating health facilities, and to the Chókwè district authorities. We acknowledge the institutional support of the Instituto Nacional de Saúde-INS, the Institute of Tropical Medicine-ITM, and the Chókwè Health Research and Training Centre (CITSC).

