## Supplementary materials for "Prevalence of *dhfr-dhps* sextuple mutants and gametocyte-harboring quintuple mutants resistant to sulfadoxine-pyrimethamine among pregnant women in Mozambique"

#### S1 METHODS

##### *Pfdhfr and pfdhps Genotyping - Restriction fragment length polymorphism*

We analyzed single nucleotide polymorphisms at the *Pfdhfr* (N51I, C59R, S108N and I164L) and *Pfdhps* (S436F, A437G, K540E and A581G) loci by PCR-restriction fragment length polymorphism (PCR-RFLP) using primers and nested-PCR protocols described elsewhere [1,2], with minor modifications. PCR reactions were run on a Biometra T professional gradient Thermocycler (Thistle Scientific Ltd, UK). For the *dhfr* loci, primary PCR was performed in a 25 µl mixture containing 2 µl of DNA sample (or 5 µl for samples with a parasite density below 1 parasite/µl), 1.5 mM MgCl2, 10x PCR Buffer, 200 µM dNTP, 0.2 µl of HotStarTaq DNA polymerase and 0.2 µM of each primer. A 720-base pair (bp) sequence was amplified with primer pairs AMP1/AMP2 (Table S1) with the following settings: initial denaturation 94°C for 3 min, 45 cycles of 94°C for 30 s, 45°C for 45 s and 72°C for 45 s with a final extension of 72°C for 5 min. For the nested PCR, 2 µl of primary PCR product was added to each of two PCR mixtures to create a final volume of 50 µl containing 1.5 mM MgCl2, 10x PCR Buffer, 200 µM dNTP, 0.2 µl of HotStarTaq DNA polymerase, and 0.25 µM of each primer. The primer pairs F-M4 and M3-F/ were used to amplify codon C59R and codons N51I, S108N and I164L, respectively with the following settings: initial denaturation 94°C for 3 min; 5 cycles of 94°C for 1 min, 45°C for 2 min, and 72°C for 1 min; 35 cycles of 94°C for 1 min, 45°C for 1 min, and 72°C for 1 min; and a final extension of 72°C for 10 min.

For the *dhps* loci, a primary PCR was performed in a 50 µl mixture containing 5 µl of DNA sample, 1.5 mM MgCl2, 10x PCR Buffer, 200 µM dNTP, 0.2 µl of HotStarTaq DNA Polymerase and 0.2 µM of each primer. A 710-bp sequence was amplified with primer pairs R2/R (Table S1) with the following settings: initial denaturation 94°C for 3 min; 45 cycles of 94°C for 1 min, 45°C for 45 s, and 72°C for 1 min; and a final extension of 72°C for 5 min. For the nested PCR, 2 µl of primary PCR product was used in a final volume of 50 µl containing PCR mixture with 1.5 mM MgCl2, 10x PCR Buffer, 200 µM dNTP, 0.2 µl of HotStarTaq DNA polymerase, and 0.2 µM of each primer pair. The following primer pairs were used: K-K/ for A437G, and K540E, J-K for S436F, and L-L/ for A581G and A613S. The settings were: initial denaturation 94°C for 2 min; 5 cycles of 94°C for 2 min, 45°C for 2 min, and 72°C for 1.5 min; 35 cycles of 94°C for 1 min, 45°C for 1 min, and 72°C for 1 min; and a final extension of 72°C for 10 min.

Five microliters (100-250 ng) of the nested PCR products containing the polymorphic regions were subjected to enzymatic digestion to detect mutations at the various sites. Single enzymatic digestions were conducted in a final volume reaction of 15 µl following the manufacturer's instructions (New England Biolabs, Beverly, MA). Double enzymatic digestions at 581 codon were conducted in a final volume reaction of 50 µl. The first incubation was performed at 55°C for 5-15 min with digestion of five microliters (100-250 ng) of DNA in 10 units of BslI. Secondly, 10 units of BstUI were added into the reaction and incubated at 60°C for 5-15 min. Furthermore, double enzymatic digestions at 164 codon were conducted in a final volume reaction of 10 µl. The first incubation was performed at 37°C for 1.5 hours with digestion of five microliters (100-250 ng) of DNA in five units of DraI. Secondly, 10 units of DraI were added into the reaction and incubated at 37°C for 1.5 hours followed by enzyme inactivation at 65°C for 20 min.

Primer sequences, restriction enzyme digestion, and fragment sizes are shown in Table S1. Plasmid controls for PCR-RFLP were obtained from MR4/BEI resources (https://www.beiresources.org/MR4Home.aspx) and were used as wild-type and mutant controls for *pfdhfr* and *pfdhps* polymorphisms. Specifically: i) FR-3D7 (MRA199) and FR-V1/S (MRA195) used as wild-type and mutant controls for *dhfr* N51I, C59R, S108N, I164L, respectively; ii) PS-FCR (MRA192) used for *dhps* S436F and A437G wild-types; iii) PS-Dd2 (MRA193) used for *dhps*

S436F mutant; iv) PS-Mali (MRA191) used for *dhps*, K540E, A581G wild-type; and v) PS-Peru (MRA190) used for *dhps*, A437G, K540E, A581G mutants. These materials were contributed by MR4/BEI Resources, NIAID, NIH, and were distributed by BEI Resources, NIAID, NIH. The use of these materials does not imply endorsement by the National Institutes of Health. The digested products were analyzed by 2% agarose (Eurogentec SA) gel electrophoresis and visualized by ultraviolet transilluminator after staining with ethidium bromide. Negative (water and human-negative DNA) controls were included in each experiment.

***Reverse transcription quantitative PCR for detection of Pfs25 gametocyte-specific transcripts***

One hundred µl of whole blood collected into 500 µl of RNAprotect stabilizer reagent (Qiagen, Germany) were used to extract RNA using RNeasy Plus 96 Kit (Qiagen) and eluted in 45 µl of the RNA Storage Solution (Ambion). Reverse transcription quantitative PCR (RT-qPCR) were performed as previously described [3]. Twenty µl reaction mixtures were run in a LightCycler® 480 Instrument (Roche Molecular Systems, Inc) using the LightCycler MultiPlex RNA virus Master kit (Roche, 7083173001) with primers and HEX-BHQ1 labelled hydrolysis probe (250 mM, IDT) published by Wampfler *et al*., (2013) [4]. All samples were tested in duplicate reactions and controls without reverse transcriptase (RT) enzyme were added to exclude false positives due to the presence of genomic DNA. The analysis was run using LightCycler480 software version 1.5.0. *P. falciparum* gametocyte densities were quantified using a 7-point standard curve ranging from $10^5$ to 0.1 gametocytes/µl generated from cultured 3D7 *P. falciparum* stage V gametocytes as described elsewhere [3]. The limit of detection (LOD) of this assay was 0.1 gametocytes/µl. Samples with Ct values higher than the last standard curve point, were considered to contain $< 0.1$ gametocytes/µl, *i.e.,* LOD.

### S2 TABLES

**Table S1.** Primer pairs and restriction enzymes used for *pfdhfr* and *pfdhps* polymorphism detection.

| Primer name | Primer sequence | Size of amplicon (bp) | Targeted mutation | Restriction enzyme | Fragment length (bp) Wild Type | Fragment length (bp) Mutant |
|---|---|---|---|---|---|---|
| **AMP1/ AMP2** | 5'-TTTATATTTTCTCCTTTTTA-3'<br>5'-CATTTTATTCGTTTTCT-3' | 720 | | | | |
| M3-F/ | 5'- TTTATGATGGAACAAGTCTGCGACGTT- 3''<br>5' - AAATTCTTGATAAACAACGGAACCTttTA- 3' | 522 | N51I | *Mlu*CI | 55, 65, 120, 153 | 55, 65, 120, 218 |
| F-M4 | 5′-GAAATGTAATTCCCTAGATATGGAATATT-3′<br>5′-TTAATTTCCCAAGTAAAACTATTAGAGCTTC-3′ | 326 | C59R | *Xmn*I | 189, 137 | 26, 137, 163 |
| M3-F/ | 5'- TTTATGATGGAACAAGTCTGCGACGTT- 3''<br>5' - AAATTCTTGATAAACAACGGAACCTttTA- 3' | 522 | S108N | *Bsr*I | 522 | 190, 332 |
| M3-F/ | 5'- TTTATGATGGAACAAGTCTGCGACGTT- 3''<br>5' - AAATTCTTGATAAACAACGGAACCTttTA- 3' | 522 | I164L | *Dra*I | 107, 171, 245 | 107, 143, 245 |
| **R2/R** | 5'- AACCTAAACGTGCTGTTCAA- 3'<br>5' - AATTGTGTGATTTGTCCACAA-3' | 710 | | | | |
| J-K | 5’- TGCTAGTGTTATAGATATAGGatGAGcATC-3'<br>5’ – CTATAACGAGGTATTgCATTTAATgCAAGAA- 3' | 438 | S436F | *MspA*1I | 438 | 32, 406 |
| K-K/ | 5’- TGCTAGTGTTATAGATATAGGatGAGcATC-3'<br>5’ – CTATAACGAGGTATTgCATTTAATgCAAGAA- 3' | 438 | A437G | *Ava*II | 438 | 34, 404 |
| K-K/ | 5’- TGCTAGTGTTATAGATATAGGatGAGcATC-3'<br>5’ – CTATAACGAGGTATTgCATTTAATgCAAGAA- 3' | 438 | K540E | *Fok* I | 33, 405 | 85, 320 |
| L-L/‡ | 5' - ATAGGATACTATTTGATATTGGAccAGGATTcG-3' | 161<br>161 | A581G | *Bst*UI and *BslI** | 105, 33, 23 | 138, 23 |

**Table S2.** Prevalence of *P. falciparum dhfr* and *dhps* haplotype combinations found at a low frequency.

| **Haplotype type** | Mutation alleles | *n* (prevalence) |
|---|---|---|
| ***dhfr*** | 51I/C59/108N/I164 | 5 (5%) |
| | 51I/59R/S108/I1S1S1S164 | 1 (1%) |
| | 51I/C59/S108/I164 | 1 (1%) |
| | N51/59R/108N/I164 | 3 (3%) |
| ***dhfr/dhps*** | 108N+540E/581G | 1 (1%) |
| | 108N+163S | 1 (1%) |
| | 108N+163S | 1 (1%) |
| | 108N+WT | 1 (1%) |
| | 108N+437G/540E/581G | 1 (1%) |
| | WT+437G/581G | 1 (1%) |
| | 59R/108N+WT | 7 (7%) |
| | 59R/108N+437G/540E | 1 (1%) |
| | 59R/108N+540E | 5 (5%) |
| | 59R/108N+581G | 1 (1%) |
| | 59R/108N+436F/437G | 1 (1%) |
| | 59R/108N+540E/163S | 1 (1%) |
| | 59R/108N/164L+540E | 1 (1%) |
| | 59R/108N/164L+437G/163S | 1 (1%) |
| | 59R+437G/540E | 1 (1%) |
| | WT+WT | 2 (2%) |
| | WT+437G/540E | 1 (1%) |
| | WT+437G | 1 (1%) |
| | WT+581G | 1 (1%) |
| | WT+437G/540E/581G | 1 (1%) |
| | WT+540E | 1 (1%) |
| | WT+437G | 1 (1%) |
| | 59R/108N+437G/540E | 1 (1%) |
| | 59R/108N+WT | 1 (1%) |
| | 59R/108N+437G/540E/581G | 1 (1%) |

**Abbreviations:** *dhfr,* dihydrofolate reductase gene; *dhps*, dihydropteroate synthetase gene; WT, wild-type.

**Table S3.** Univariate analysis of risk factors associated with carriage of quintuple and sextuple mutant haplotypes (*n* = 100).

| | | **Quintuple haplotype (*n* = 54) §** | | | | **Sextuple haplotype (*n* = 7) ‡** | | | |
|---|---|---|---|---|---|---|---|---|---|
| **Variable** | ***n*** | ***n* [%]** | **OR** | **95% CI** | **p-*value*** | ***n* [%]** | **OR** | **95%CI** | **p-value** |
| Age (years) | | | | | | | | | |
| < 20 | 40 | 21 (52.5) | 0.9 | 0.4-2.0 | 0.81 | 1 (2.5) | 0.2 | 0.01-1.4 | 0.18 |
| ≥ 20 | 60 | 33 (61.1) | | Ref | | 6 (10.0) | | Ref | |
| *Education* | | | | | | | | | |
| None/primary | 53 | 35 (66.0) | **2.9** | **1.3-6.6** | **0.01** | 4 (7.6) | 1.2 | 0.3-6.3 | 0.82 |
| Secondary | 47 | 19 (40.4) | | Ref | | 3 (6.4) | | Ref | |
| *Place of residence* | | | | | | | | | |
| Urban | 57 | 26 (45.6) | | Ref | | 6 (10.5) | | Ref | |
| Rural | 43 | 28 (65.1) | **2.2** | **0.9-5.0** | **0.05** | 1 (2.3) | 0.2 | 0.01-1.3 | 0.15 |
| *Gravidity* | | | | | | | | | |
| Primigravidae (1) | 45 | 22 (48.8) | 0.7 | 0.3-1.5 | 0.35 | 2 (4.4) | 0.5 | 0.1-2.3 | 0.38 |
| Multigravidae (≥2) | 55 | 31 (58.2) | | Ref | | 5 (9.1) | | Ref | |
| *No. IPTp-SP doses received* | | | | | | | | | |
| < 3 doses | 46 | 27 (58.7) | | Ref | | 3 (6.5) | | Ref | |
| ≥ 3 doses | 54 | 27 (50.0) | 1.4 | 0.6-3.2 | 0.39 | 4 (7.4) | 0.9 | 0.2-4.1 | 0.86 |
| *Parasite density, p/µl***Ψ** | | | | | | | | | |
| <100 p/µL | 67 | 37 (55.2) | | Ref | | 5 (7.5) | | Ref | |
| ≥100 p/µL | 33 | 17 (51.5) | 0.9 | 0.9-1.9 | 0.73 | 2 (6.1) | 0.8 | 0.1-4.4 | 0.77 |

**Ψ** Peripheral parasite density

**§** Adjusted for age, education, residence location, gravidity, IPTp uptake and parasite density

**‡** Adjusted for education, gravidity, IPTp uptake and parasite density

**Boldface** indicates statistical significance ($p < 0.05$).

**Abbreviations:** CI, confidence interval, IPTp-SP, intermittent preventive treatment for malaria in pregnancy with sulfadoxine-pyrimethamine, OR, odds ratio.

**Table S4.** Effect of mutant haplotypes and other risk factors on adverse pregnancy outcomes in Chókwè district (*n* = 100) * (Univariate analysis).

| **Variable** | ***n*** | **BW < 2500g** | | | **Placental malaria ¥** | | | **Gestational age** (<37wk)** | | |
|---|---|---|---|---|---|---|---|---|---|---|
| | | ***n* [%]** | **OR [95%CI]** | ***p-value*** | ***n* [%]** | **OR [95%CI]** | ***p-value*** | ***n* [%]** | **OR [95%CI]** | ***p-value*** |
| Mutant haplotype | | | | | | | | | | |
| Sextuple‡ | 7 | 1 (14.3) | 1.4 (0.1-15.3) | 0.75 | 1 (14.3) | 0.3 (0.03-2.7) | 0.28 | 2 (28.6) | 1.2(0.2-6.9) | 0.87 |
| Quintuple § | 54 | 3 (5.6) | 0.52 (0.1-2.4) | 0.40 | 13 (24.1) | 0.6 (0.2-1.3) | 0.21 | 9 (16.7) | 0.5 (0.2-1.6) | 0.29 |
| Others¶ | 39 | 4 (10.3) | Ref | | 14 (35.9) | Ref | | 10 (25.6) | Ref | |
| Age (years) | | | | | | | | | | |
| < 20 | 40 | 6 (15) | **5.1(0.9-26.7)** | **0.05** | 13 (32.5) | 1.4(0.6-3.5) | 0.41 | 7 (17.5) | 0.7(0.3-1.9) | 0.48 |
| ≥ 20 | 60 | 2 (3.3) | Ref | | 15 (25.0) | Ref | | 14 (23.3) | Ref | |
| *Education* | | | | | | | | | | |
| None/primary | 53 | 4 (3.7 | 0.8(0.2-4.8) | 0.85 | 13 (24.5) | 0.7(0.3-1.7) | 0.41 | 10 (18.9) | 0.7(0.3-1.) | 0.56 |
| Secondary | 47 | 4 (8.5) | Ref | | 15 (31.9) | Ref | | 11 (23.4) | Ref | |
| *Place of residence* | | | | | | | | | | |
| Urban | 57 | 5 (8.8 | Ref | | 18 (31.8) | Ref | | 13 (24.1) | Ref | |
| Rural | 43 | 3 (6.9 | 0.78(0.2-3.4) | 0.74 | 10 (23.3) | 0.6(0.3-1.6) | 0.36 | 8 (18.6) | 0.7(0.3-2.1) | 0.61 |
| *Gravidity* | | | | | | | | | | |
| Primigravidae (1) | 45 | 8 (17.7) | N/A£ | N/A | 14 (31.1) | 1.4(0.5-3.9) | 0.54 | 6 (13.3) | 0.4(0.1-1.2) | 0.09 |
| Multigravidae(≥ 2) | 55 | 0 (0.0) | | | 14 (25.5) | Ref | | 15 (27.3) | Ref | |
| *No. IPTp doses received* | | | | | | | | | | |
| < 3 doses | 46 | 1 (2.2) | 0.1(0.2-1.3) | 0.08 | 13 (28.3) | 1.0(0.4-2.5) | 0.96 | 6 (13.0) | 0.4(0.1-1.1) | 0.08 |
| ≥ 3 doses | 54 | 7 (13.0) | Ref | | 15 (27.8) | Ref | | 15 (27.8) | Ref | |

**Boldface** indicates statistical significance ($p < 0.05$). **¥** placental malaria as detected by histology, **£** analysis not done for birth weight due to observations in multigravidae. ** indicates gestational age at delivery, ***§*** (**IRN**-**GE**, **ǂ IRN**-**GEG, ¶** include WT and other mutant combinations**. Abbreviations:** BW, birth weight; CI, confidence interval; wk, weeks; IPTp-SP, intermittent preventive treatment for malaria in pregnancy with sulfadoxine-pyrimethamine; OR, odds ratio.

**Table S5.** Parasite variability shown as Multiplicity of infection (MOI) in different mutation haplotypes

| Variable | N | Median [IQR] | Type of infection | | *p*-value |
|---|---|---|---|---|---|
| | | | Monoclonal (MOI = 1) | Polyclonal (MOI ≥ 2 ) | |
| **Mutations in participant** | | | | | |
| Wild-type | 12 | 4.5 [2.5-5.5] | 1 | 11 | 0.63 |
| Any mutation | 83 | 3 [2.0-4.0] | 11 | 72 | |
| **Mutant haplotype‡** | | | | | |
| Sextuple | 5 | 3 [2.0-3.0] | 0 | 5 | |
| Quintuple | 54 | 3 [2.0-3.0] | 10 | 44 | 0.13 |
| Other* | 36 | 3.5 [2.5-5.0] | 2 | 34 | |

*Composition of other mutations and combinations is presented in Table S2. **Abbreviations:** IQR, Interquartile range.

**‡** two samples were excluded from the analysis due to inconclusive results for MOI assay.

**Table S6.** Frequency of gametocyte carriage and gametocyte densities by IPTp-SP uptake group.

| | **IPTp-SP dose** | | | | | |
|---|---|---|---|---|---|---|
| | **Total** | **None** | **One** | **Two** | **≥ 3** | |
| | **N = 100** | ***n* = 12** | ***n* = 11** | ***n* = 23** | ***n* = 54** | ***p*-value** |
| Gametocyte carrier | 34 | 6 | 5 | 6 | 17 | |
| % (n/N) | 34.0 | 50.0 | 45.5 | 26.1 | 31.5 | 0.40 |
| Median gametocyte density (n=15) * | 1.76 | 0.66 | 0.15 | 1.86 | 1.87 | 0.39 |
| Interquartile range [IQR] | [0.62-3.19] | [0.62-2.06] | [0.15-0.15] | [0.64-3.54] | [1.75-5.57] | |

Gametocyte carriage was defined as presence of gametocytes detected by RTqPCR. Gametocyte densities were quantified by RT-qPCR, log10-transformed for regression analyses and raw values (per µl) are reported in this analysis. *Only 15 (44.1% [15/34]) samples could be quantified; 19 (55.9% [19/34]) had a gametocyte density of <0.1 gametocytes/µl. **Statistical tests** used to assess differences between IPTp-SP uptake groups: Chi-square ($\chi^2$) test (group counts > 5) for gametocyte carriage, and Kruskal-Wallis test for gametocyte densities. **Abbreviations and definitions:** IPTp-SP, Intermittent preventive treatment in pregnancy with Sulfadoxine-Pyrimethamine; RT-qPCR, reverse transcription quantitative polymerase chain reaction
